# Natural History of Accessory Navicular in Children: A Longitudinal Study with 2-Year Follow-up – KID Locomo Study

**DOI:** 10.1101/2025.07.02.25330707

**Authors:** Takahide Sasaki, Masatoshi Teraguchi, Kanae Mure, Yoshiki Asai, Yusuke Kido, Makiko Onishi, Kazuyoshi Minamino, Takashi Shimoe, Nobuyuki Miyai, Yukihiro Nakagawa, Hiroshi Hashizume, Hiroshi Yamada

## Abstract

**Introduction:** The accessory navicular (AN) is an accessory ossicle located on the medial side of the navicular bone and is often associated with sports-related overuse injuries during adolescence. However, little is known about the natural history of AN. This study aimed to clarify the natural course of AN, including symptomatic cases, in children through a longitudinal epidemiological investigation.

**Methods:** Data from the KID Locomo Study, a prospective cohort study aimed at elucidating musculoskeletal disorders in childhood, were used. Of the 834 children recruited in the 2022 baseline survey, 66 children (109 feet) aged 11 to 13 years with AN were enrolled in this 2-year longitudinal analysis. The presence of AN was assessed using ultrasonography. Data on sex, age, height, weight, presence of pain at the AN site, laterality, flatfoot (based on footprint analysis), and participation in athletic clubs were collected. The natural history of AN, including painful presentations, was evaluated. Multivariable logistic regression analysis was performed to identify risk factors for non-fusion of the AN with the navicular bone.

**Results:** Among the 109 feet with AN at baseline, 38 (34.9%) demonstrated fusion with the navicular bone at the 2-year follow-up. Of the 21 feet with painful AN at baseline, pain had resolved in 15 (71.4%) after 2 years. Multivariable logistic regression analysis revealed that female sex (odds ratio: 3.00; 95% confidence interval: 1.26–7.45; p = 0.01) and higher body mass index (BMI) (odds ratio: 1.20; 95% confidence interval: 1.02–1.43; p = 0.02) were significant risk factors for non-fusion.

**Conclusions:** This study provides epidemiological data on the natural history of AN, including symptomatic cases, in children. To our knowledge, this is the first study to report the natural course of painful AN. These findings may offer fundamental insights into prevention and management strategies for painful AN in children.

## Introduction

The accessory navicular (AN) is an accessory bone located on the posteromedial side of the navicular tuberosity, originating from a developmental anomaly of the secondary ossification center (1). It is a common cause of medial foot pain in adolescents, particularly in those engaged in athletic activities (2) (3). Treatment of painful AN is typically categorized into conservative and surgical approaches (1) (4). Conservative management includes rest, cast immobilization, physical therapy, oral nonsteroidal anti-inflammatory drugs, and orthotics (5) (6). In cases unresponsive to conservative treatment, surgical options such as excision, percutaneous drilling, or the Kidner procedure may be indicated (4) (7) (8) (9).

Several cross-sectional epidemiological studies have reported that the prevalence of AN ranges from 4% to 21% (7) (10) (11) (12) (13). These studies have contributed valuable prevalence data but are limited in their ability to assess time-dependent changes. Regarding its natural history, Knapik et al. reviewed 2,620 annual unilateral foot radiographs obtained from 261 pediatric participants and identified AN in 19 cases, among whom 42% (8 cases) demonstrated fusion with the navicular (5). To the best of our knowledge, this is the only longitudinal epidemiological study on AN. Although the study by Knapik et al. provides valuable insight into the longitudinal radiographic behavior of AN, including the timing of its appearance and fusion, the small number of AN cases limits its broader applicability, and the natural history of AN remains poorly understood. Moreover, no longitudinal studies have investigated symptomatic AN, and its natural course remains entirely unclarified.

This study aimed to clarify the natural history of AN and the natural course of symptomatic AN in Japanese children through a longitudinal epidemiological investigation of a general pediatric population. These findings may offer foundational insights to inform future prevention and treatment strategies for painful AN.

## Materials and Methods

### Study Design

This longitudinal study examined the natural history of AN in Japanese children. The study complied with the principles of the Declaration of Helsinki and was approved by the Ethics Committee of Wakayama Medical University (No. 3594). Written informed consent was obtained from the parents or guardians of all participants.

### Participants

The Katsuragi Integrated Defense for Locomotive Syndrome in Children Study (KID Locomo Study) is a prospective cohort study aimed at elucidating musculoskeletal disorders in childhood, identifying risk factors, and establishing preventive methods. Study participants included children from five elementary schools and two middle schools in Katsuragi Town, Wakayama Prefecture. A baseline survey was conducted in each elementary and middle school between October 2022 and February 2023. One small elementary school was surveyed together with the other schools because of its small student population. Annual follow-up surveys were conducted. A team of orthopedic surgeons, nurses, physical therapists, university faculty, and graduate students collected the data. Participants received a data collection manual and attended rehearsal sessions to standardize the protocol.

This study employed a two-year longitudinal design using data from the first and third visits of the KID Locomo Study, which served as the baseline and follow-up assessments, respectively. The baseline survey was conducted from October 4, 2022 to February 10 2023, and the follow-up from September 17, 2024 to December 13, 2024. A total of 834 children (1,668 feet; 426 males and 408 females, aged 6–15 years) participated in the baseline survey. Of these, 72.3% (603 children; 313 males and 290 females, aged 6–13 years) were successfully followed up in the third visit. Since the KID Locomo Study targeted elementary and middle school students, participants who were in the second or third year of middle school at the time of the baseline survey had graduated by the two-year follow-up and were therefore excluded from the follow-up assessment. Of the 834 children in the baseline survey, 195 (293 feet; 88 males with 131 feet and 107 females with 162 feet) were identified as having AN based on ultrasonography and were recruited for the study. Follow-up data were obtained from 135 children (202 feet; 61 males and 74 females, aged 6–13 years). In addition, 69 children aged 6–10 years at baseline were excluded. A previous study reported that the average age of appearance of AN is 12.2 ± 2.2 years in males and 10.1 ± 0.7 years in females (5). In the ultrasonographic evaluation of AN in this study, the differentiation between immature AN and normal anatomical variation was complicated in participants aged 6–10 years, as ultrasonographic imaging frequently demonstrated high-echogenic regions within the cartilaginous portion of the navicular tuberosity. As a result, 66 children (109 feet; 27 males and 39 females, aged 11–13 years) were included in the longitudinal analysis of AN. Among these, 18 children (29 feet) experienced pain in the region of the AN, and 14 children (21 feet) had follow-up assessments of pain status and were included in the analysis of the natural course of symptomatic AN (Fig. 1). The exclusion criteria were as follows: (1) medical conditions or musculoskeletal disorders that precluded physical function testing; (2) internal metal implants that precluded body composition testing; and (3) a history of foot surgery or paralysis. No participants met these criteria.

**Fig. 1.**
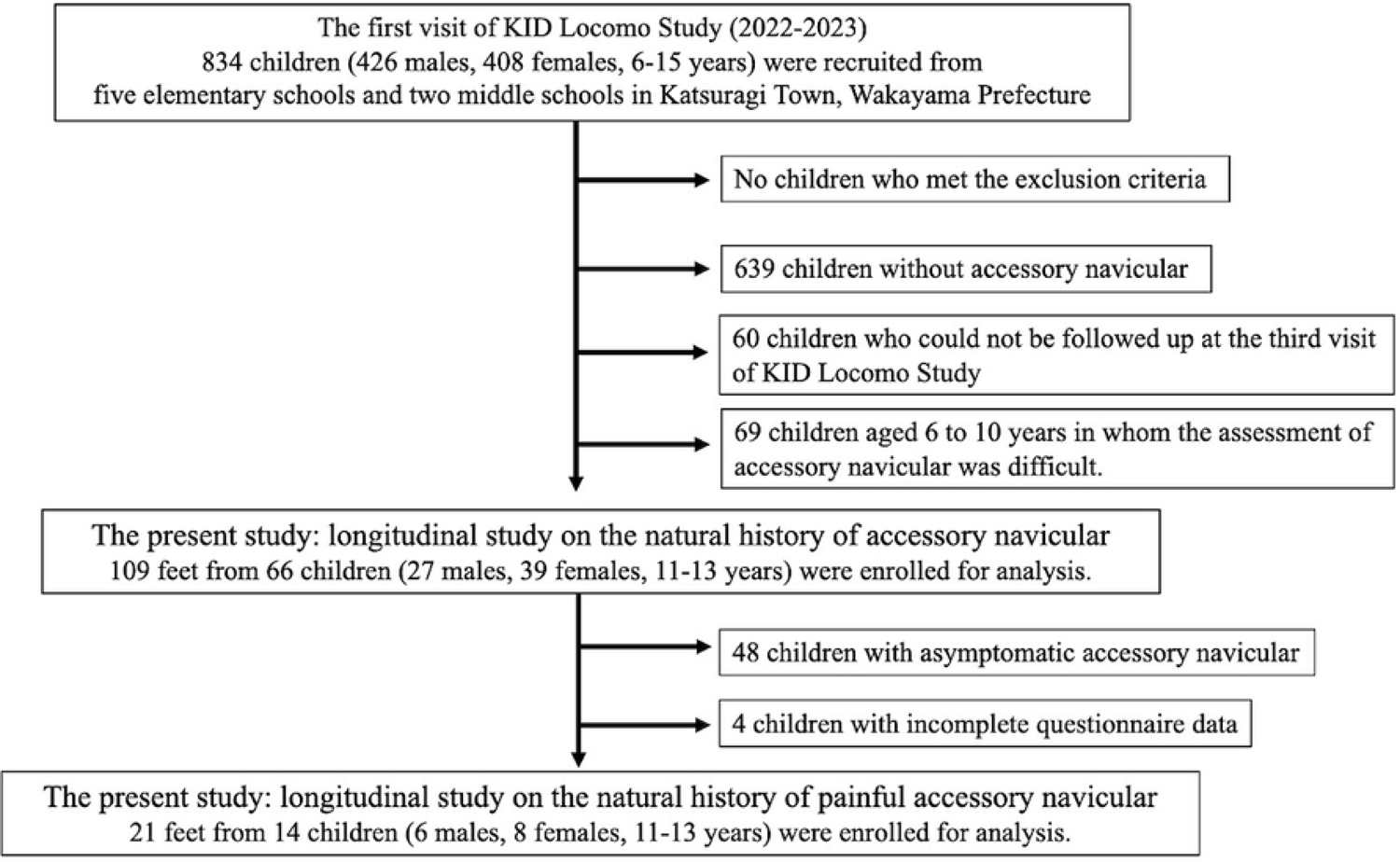
Flow diagram depicting participants recruited to the present study.

### Measurements

The KID Locomo Study collected data on: (1) height and weight; (2) physical function tests (two-step test, stand-up test, standing on one leg, squatting, arm raising, forward flexion); (3) body composition using bioelectrical impedance analysis (MC780-AN, Tanita Corp., Tokyo, Japan); (4) bone density of the calcaneus using quantitative ultrasound (Benus II, Ishikawa Seisakusho, Kanazawa, Japan); (5) posture using a computer-assisted, non-invasive device (Spinal Mouse, Idiag, Volkerswill, Switzerland); (6) fat degeneration of the quadriceps using ultrasound (an 11-MHz high-resolution linear-array transducer, SONIMAGE MX1, Konica Minolta, Tokyo, Japan); (7) presence of AN using ultrasonography; (8) presence of flatfoot based on footprint analysis (Sorbo Foot Printer, Sanshin Enterprises Co., Ltd., Tokyo, Japan); and (9) various data collected through standardized self-reported questionnaires (demographic information; lifestyle habits; exercise habits, including club activities; shoulder stiffness; lower back pain; the Strengths and Difficulties Questionnaire [SDQ] (14); and the Brief Self-Administered Diet History Questionnaire for adolescents [BDHQ-15y] (15)). Height and weight were measured using a fixed stadiometer and a digital scale, respectively, to calculate body mass index (BMI). For participants identified as having AN through ultrasonography, an interview was conducted to assess the presence of pain at the AN site (at rest, during walking, or during exercise). Due to personnel constraints, the follow-up phase did not include flatfoot assessment via footprint analysis. The present study incorporated the following variables from the baseline survey: demographic characteristics, anthropometric data, club activity status, presence of AN, associated pain, and flatfoot status. From the follow-up data, the presence of AN and pain status were analyzed. Participants with AN at baseline but not at follow-up were categorized as having fused AN, while those with persistent findings were categorized as having unfused AN.

### Evaluation of the accessory navicular using ultrasound

As this study targeted a general pediatric population, the presence of AN was assessed using ultrasonography instead of X-rays to avoid radiation exposure. The evaluation of AN via ultrasonography followed the methods reported in a previous study (16) (17). Participants were seated on a chair with slightly flexed hip and knee joints and with their feet in a frog-leg position, with the medial side facing upward on the chair. The navicular tuberosity was first palpated, and the probe was placed directly over it along the foot’s longitudinal axis. The navicular tuberosity and the attachment of the posterior tibial tendon were visualized, and the AN was identified on the proximal medial side of the navicular bone (Fig. 2). The navicular tuberosity was carefully examined from the plantar to the dorsal aspect for the presence of AN. Additionally, because a type I AN in the Veitch classification is located within the posterior tibial tendon and is separated from the navicular bone, the distal portion of the posterior tibial tendon was meticulously examined for AN (1) (4) (18). Ultrasonography was performed on both feet.

**Fig 2.**
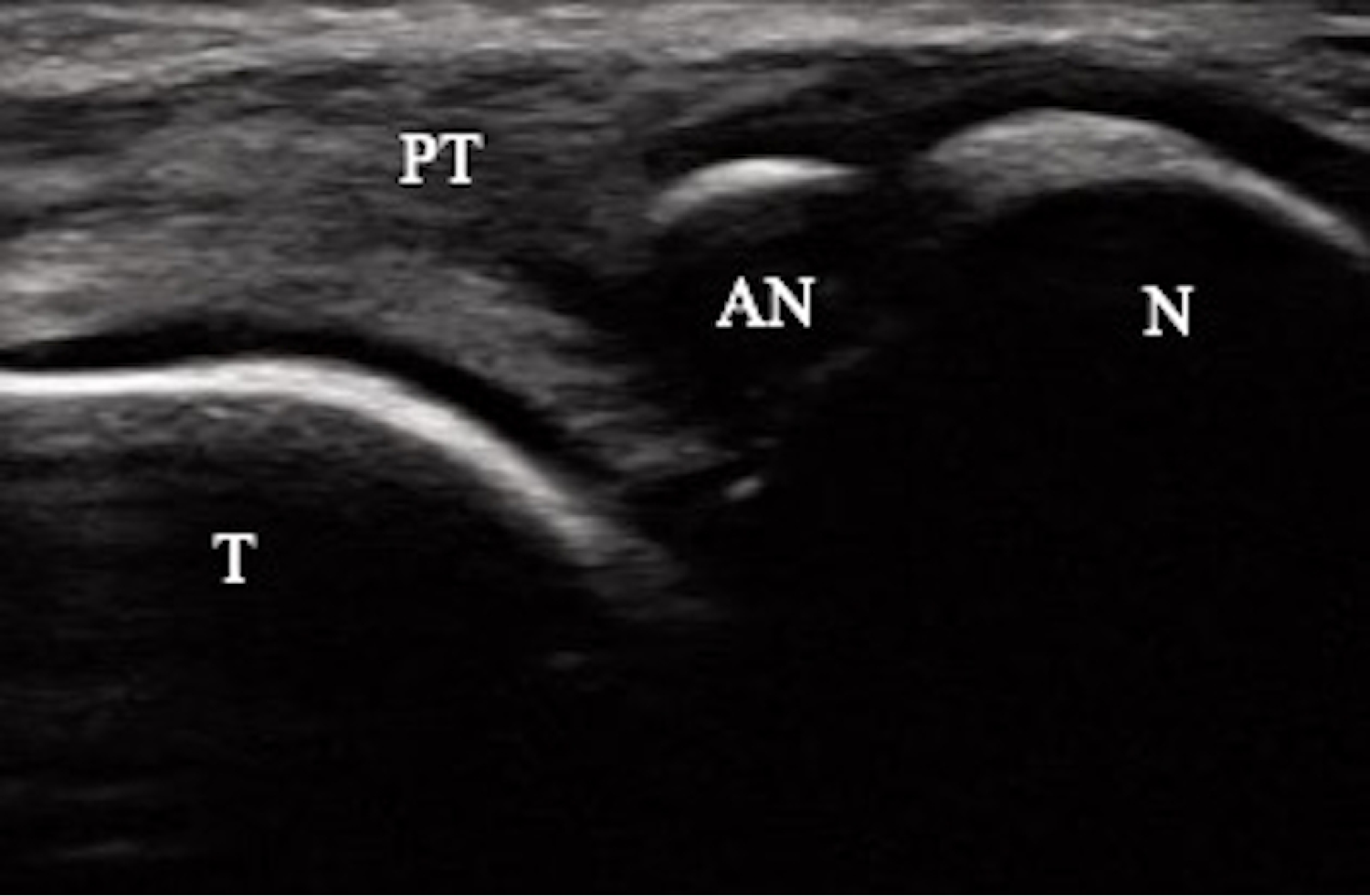
Ultrasonographic findings of the accessory navicular bone.

The accessory navicular bone is identified on the proximal side of the navicular bone. PT = posterior tibial tendon; AN = accessory navicular; N = navicular

The evaluation of AN via ultrasonography was conducted at each elementary and junior high school by two board-certified foot and ankle surgeons (TS and YA), and the results were used for analysis. The ultrasound data were saved in video format, and 80 cases were randomly selected from the stored data. To calculate intraobserver and interobserver reliability, TS and YA independently evaluated each of the 80 cases—with all participant information blinded—separately from the initial on-site evaluations. TS performed assessments twice, with a one-month interval between them, whereas YA performed a single assessment. Intraobserver reliability was calculated based on the two assessments conducted by TS. Interobserver reliability was calculated based on the first assessment by TS and the assessment by YA. A kappa value greater than 0.90 was considered excellent, and a value between 0.80 and 0.90 was considered good (19) (20). Disagreements in ultrasound image classification were resolved by consensus after the reliability assessments were completed.

### Evaluation of flatfoot using footprint analysis

Flatfoot was evaluated in this study using footprint analysis, following established protocols from previous research (21). Participants were seated and instructed to place the foot under examination on the footprint apparatus. Footprints were recorded in a full weight-bearing position, and both feet were analyzed. The Staheli Arch Index, previously validated as a reliable metric, was calculated from the obtained footprints (21) (22) (23). This index quantifies the ratio between the widths of the central (A) and heel (B) regions of the footprint. A tangential line was drawn between the medial forefoot and heel, and perpendicular lines from the midpoint and the heel contact point were used to measure A and B, respectively. An index greater than 1.0 was defined as indicative of flatfoot (Fig. 3) (22).

**Fig 3.**
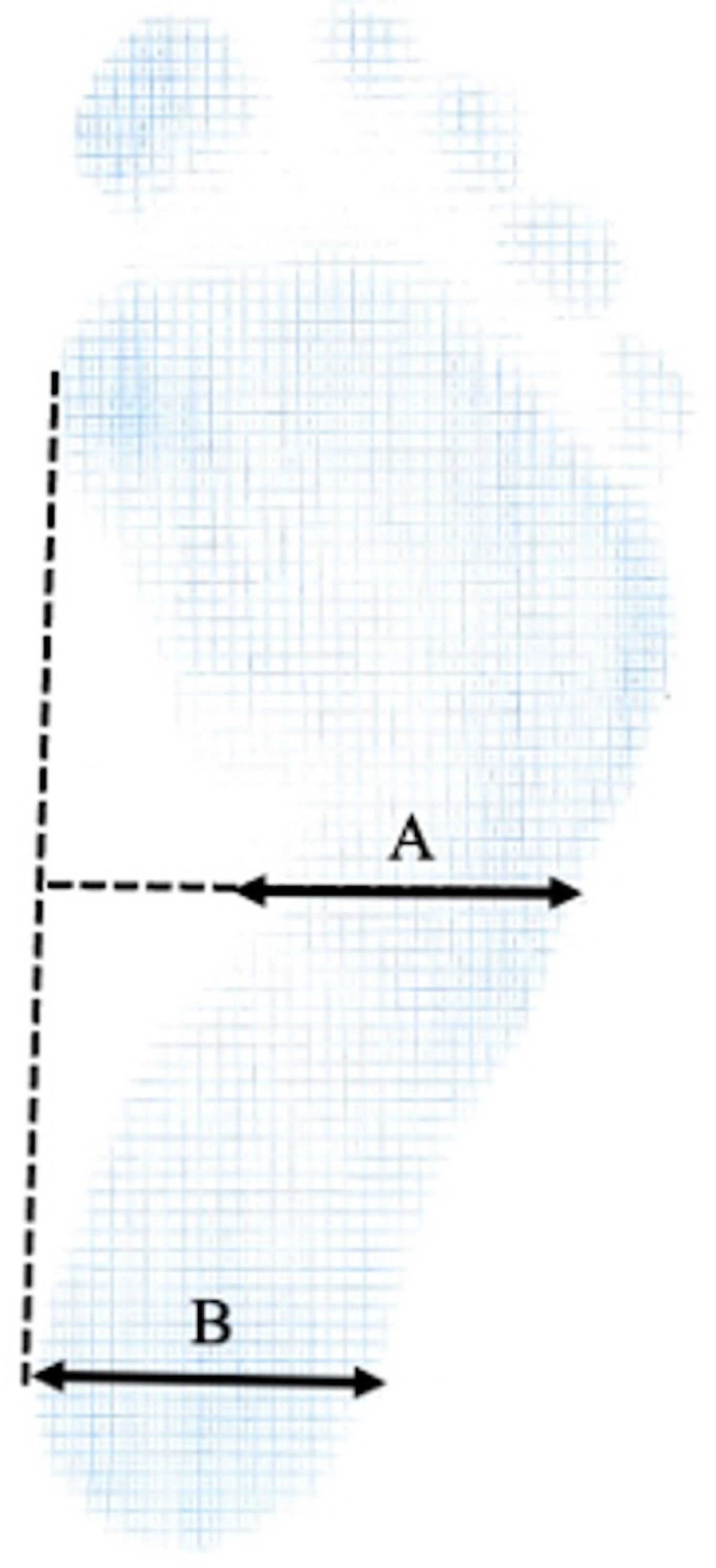
Measurement of the Staheli’s Arch Index.

To assess measurement reliability, 80 cases were randomly selected from the stored footprint data in PDF format. Two independent examiners, TS and KM, conducted the evaluations. TS performed two assessments with a one-month interval, while KM conducted a single assessment. Intraobserver reliability was determined from TS’s two evaluations, and interobserver reliability was based on the first evaluation by TS and the assessment by KM. A kappa coefficient greater than 0.90 was considered excellent, and values between 0.80 and 0.90 were considered good (19) (20). Any discrepancies were resolved through consensus following the reliability assessments.

### Statistical analyses

Baseline participant characteristics were summarized using descriptive statistics. The proportions of AN fusion and resolution of pain in symptomatic AN were calculated for the entire cohort and stratified by sex and age. Univariate analyses were conducted to examine associations between AN fusion and several variables, including sex, age, height, weight, BMI, presence of pain, flatfoot status, sports club participation, and laterality. The Wilcoxon test was used for continuous variables, and the Chi-square test was applied for categorical data. Multivariate logistic regression analysis was performed to identify risk factors for non-fusion of AN, with fusion status as the dependent variable and sex, age, BMI, flatfoot status, and sports club participation as independent variables. Due to the limited sample size, no statistical analysis was performed for factors related to the resolution of pain in symptomatic AN. All analyses were conducted based on the number of feet. Statistical analyses were performed using JMP version 14 (SAS Institute Japan, Tokyo, Japan), and a p-value less than 0.05 was considered statistically significant.

## Results

### Participant characteristics

Table 1 presents the characteristics of the 66 participants (109 feet) enrolled in this study. Of these, 41.3% were male (45 feet), and 58.7% were female (64 feet). The prevalence of symptomatic accessory navicular was 26.6% in the entire cohort (29/109 feet), 31.1% among males (14/45 feet), and 23.4% among females (15/64 feet).

**Table 1.**
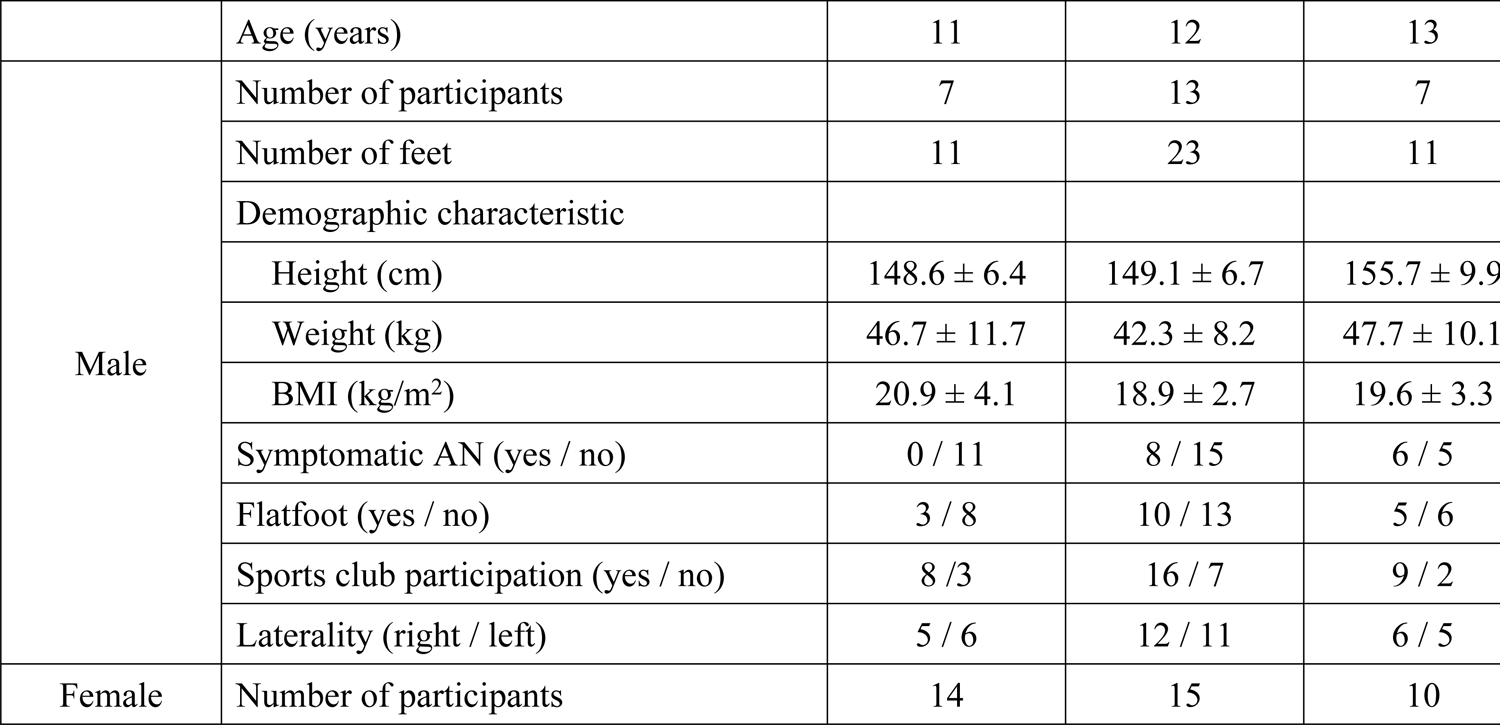
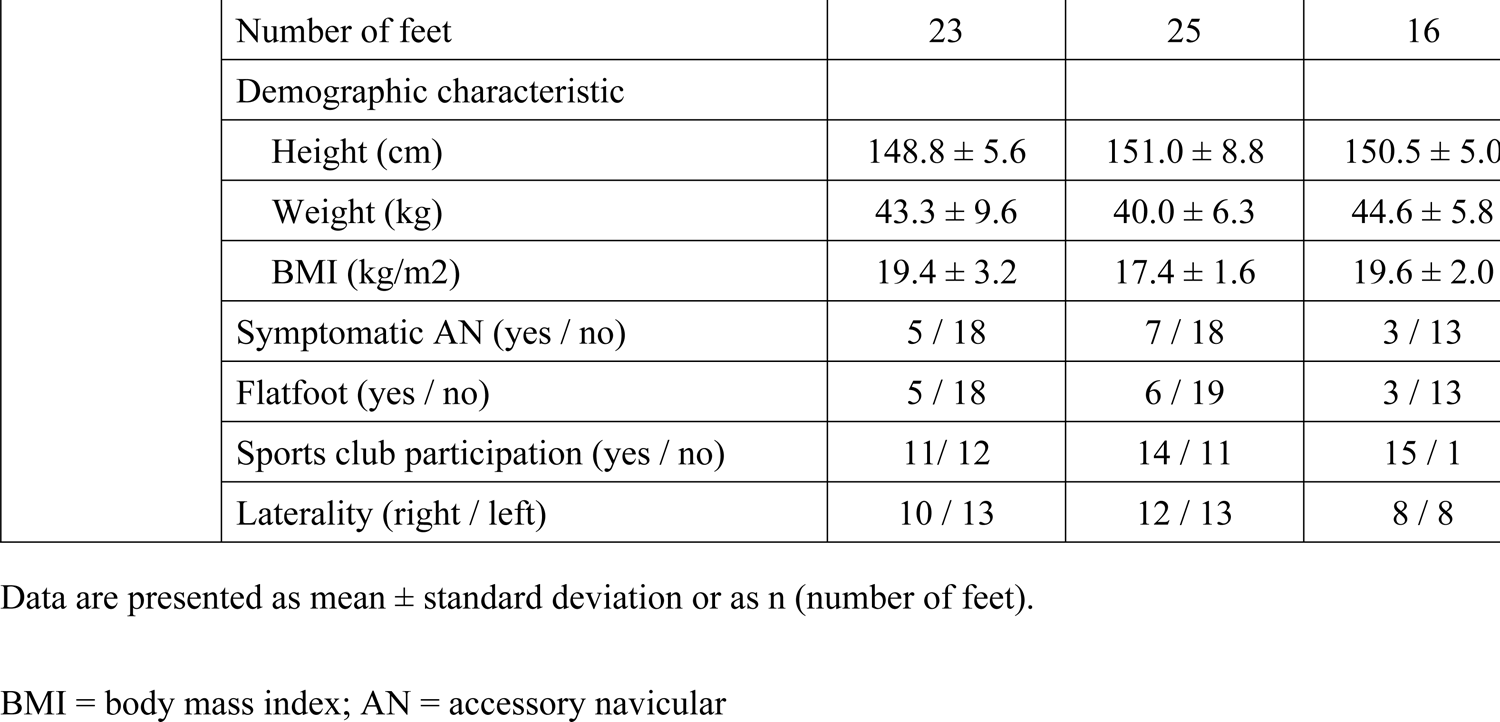
Characteristics of participants.

|  | Age (years) | 11 | 12 | 13 |
| --- | --- | --- | --- | --- |
| Male | Number of participants | 7 | 13 | 7 |
|  | Number of feet | 11 | 23 | 11 |
|  | Demographic characteristic |  |  |  |
|  | Height (cm) | 148.6 ± 6.4 | 149.1 ± 6.7 | 155.7 ± 9.9 |
|  | Weight (kg) | 46.7 ± 11.7 | 42.3 ± 8.2 | 47.7 ± 10.1 |
|  | BMI (kg/m<sup>2</sup>) | 20.9 ± 4.1 | 18.9 ± 2.7 | 19.6 ± 3.3 |
|  | Symptomatic AN (yes / no) | 0 / 11 | 8 / 15 | 6 / 5 |
|  | Flatfoot (yes / no) | 3 / 8 | 10 / 13 | 5 / 6 |
|  | Sports club participation (yes / no) | 8 / 3 | 16 / 7 | 9 / 2 |
|  | Laterality (right / left) | 5 / 6 | 12 / 11 | 6 / 5 |
| Female | Number of participants | 14 | 15 | 10 |
|  | Number of feet | 23 | 25 | 16 |
|  | Demographic characteristic |  |  |  |
|  | Height (cm) | 148.8 ± 5.6 | 151.0 ± 8.8 | 150.5 ± 5.0 |
|  | Weight (kg) | 43.3 ± 9.6 | 40.0 ± 6.3 | 44.6 ± 5.8 |
|  | BMI (kg/m2) | 19.4 ± 3.2 | 17.4 ± 1.6 | 19.6 ± 2.0 |
|  | Symptomatic AN (yes / no) | 5 / 18 | 7 / 18 | 3 / 13 |
|  | Flatfoot (yes / no) | 5 / 18 | 6 / 19 | 3 / 13 |
|  | Sports club participation (yes / no) | 11 / 12 | 14 / 11 | 15 / 1 |
|  | Laterality (right / left) | 10 / 13 | 12 / 13 | 8 / 8 |

Data are presented as mean ± standard deviation or as n (number of feet).
BMI = body mass index; AN = accessory navicular

### Natural history of accessory navicular

Among the 109 feet in which AN was identified at baseline, 65.1% (71/109 feet) still showed AN at follow-up, while 34.9% (38/109 feet) had fused with the navicular bone. Fig 4 presents the fusion rates of AN by sex and age. In males, the overall fusion rate was 46.7% (21/45 feet), with rates of 54.5% (6/11 feet) at age 11, 52.2% (12/23 feet) at age 12, and 27.3% (3/11 feet) at age 13. In females, the overall fusion rate was 26.6% (17/64 feet), with rates of 13.0% (3/23 feet) at age 11, 32.0% (8/25 feet) at age 12, and 37.5% (6/16 feet) at age 13.

**Fig 4.**
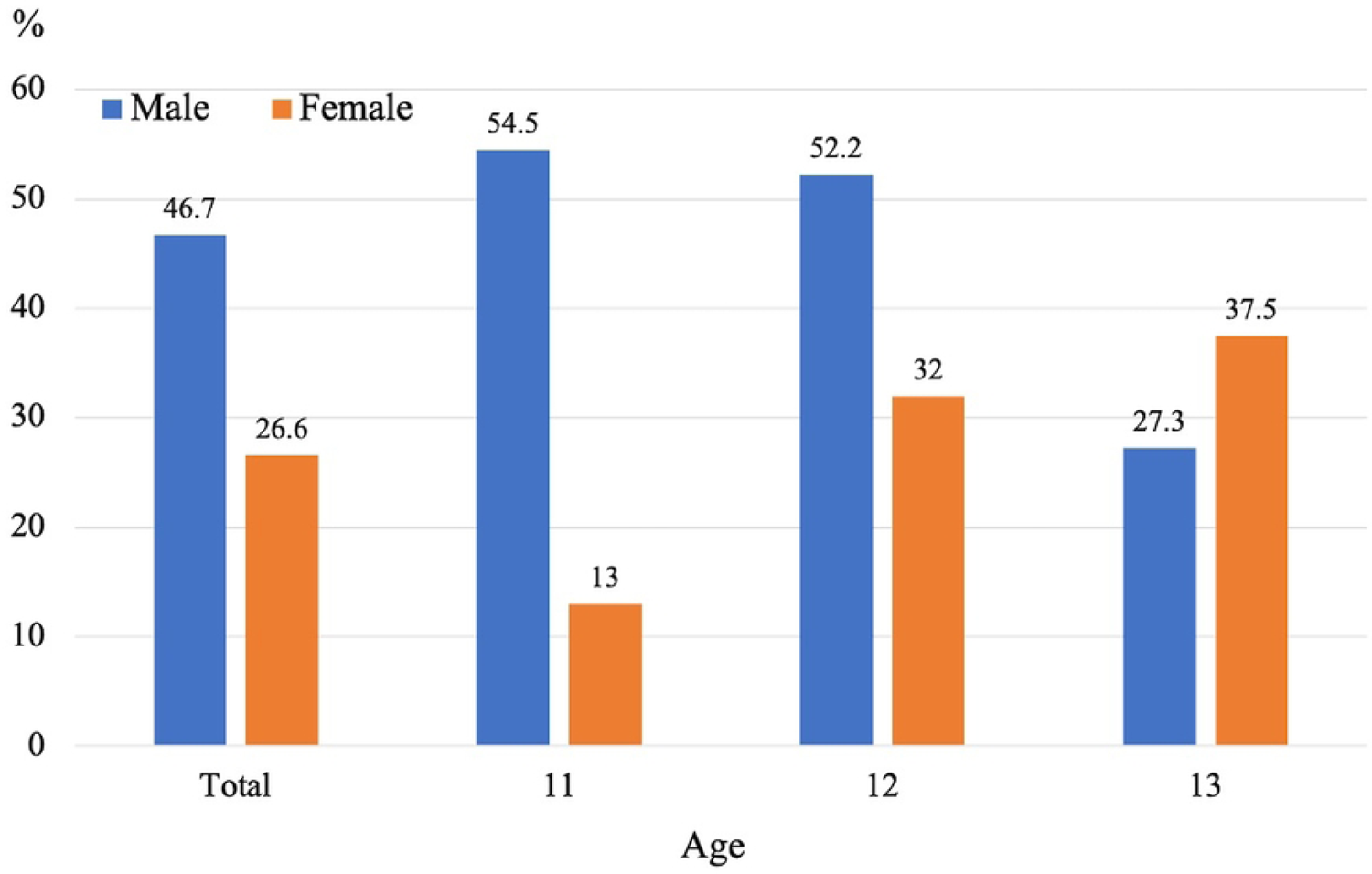
Fusion rates of accessory navicular by sex and age.

### Natural history of painful accessory navicular

Of the 29 feet identified as having a symptomatic AN at baseline, pain status could be confirmed at follow-up in 21 feet. Among these, persistent pain was observed in 28.6% (6/21 feet), while 71.4% (15/21 feet) experienced resolution of pain. All 29 feet diagnosed with symptomatic AN at baseline still exhibited AN at follow-up, indicating that no cases underwent fusion with the navicular bone. Fig 5 shows the rates of pain resolution in symptomatic AN according to sex and age. Among males, the overall rate of pain resolution was 70.0% (7/10 feet), with 66.7% (4/6 feet) in 12-year-olds and 75.0% (3/4 feet) in 13-year-olds. Among females, the overall rate was 72.7% (8/11 feet), with 50.0% (2/4 feet) in 11-year-olds and 85.7% (6/7 feet) in 12-year-olds. No male participants aged 11 years and no female participants aged 13 years presented with symptomatic AN at baseline.

**Fig 5.**
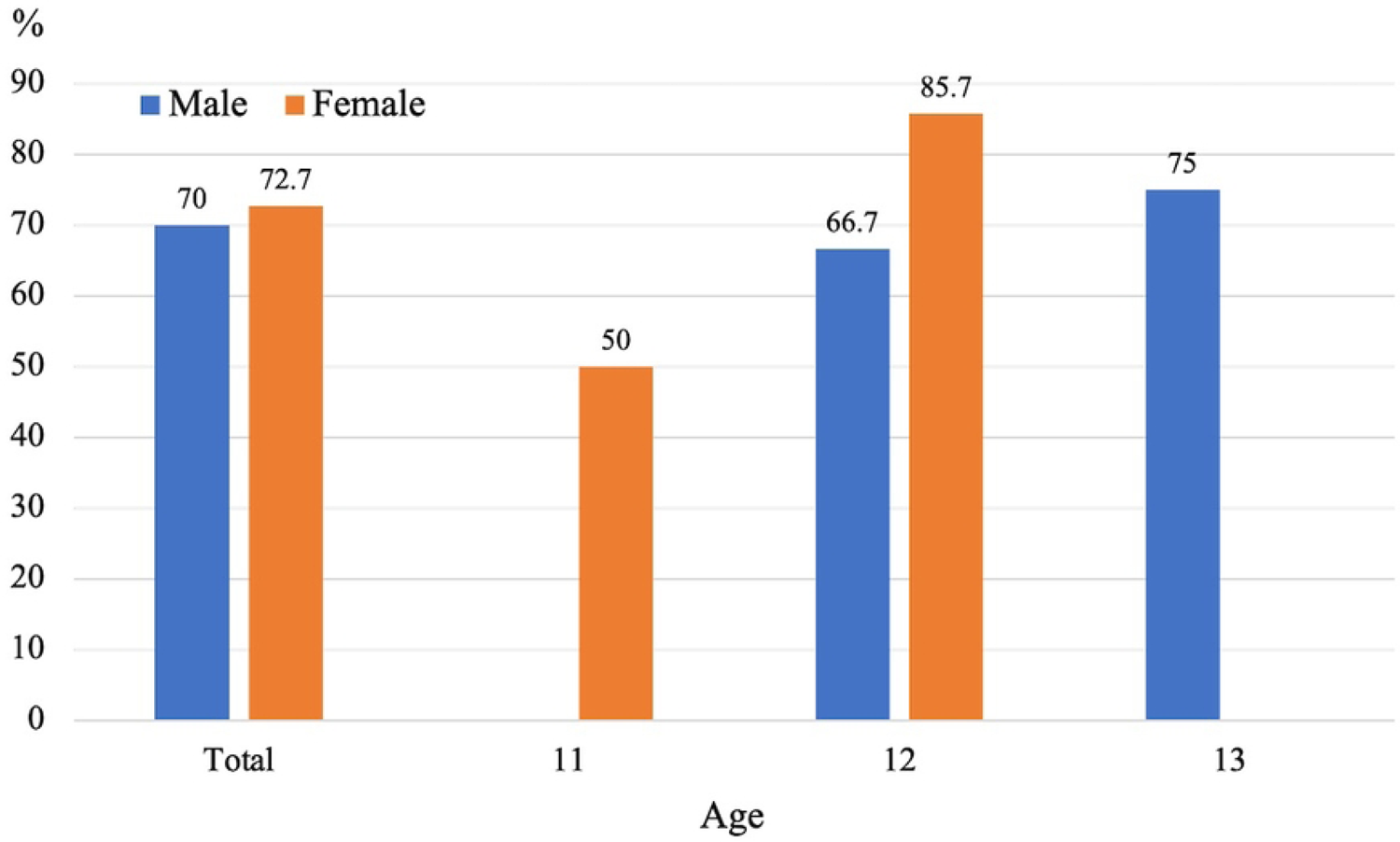
Pain resolution rates of symptomatic accessory navicular by sex and age.

### Risk factors for non-fusion of accessory navicular

Differences in demographic and clinical characteristics between participants with and without fusion of AN are shown in Table 2. The non-fusion group had a significantly higher proportion of females (p = 0.03), greater height (p = 0.003), higher body weight (p = 0.003), and higher BMI (p = 0.03) compared to the fusion group. No statistically significant differences were observed between the two groups in terms of age, presence of pain, flatfoot status, participation in athletic clubs, or laterality.

**Table 2.** Differences in demographic and clinical characteristics between participants with and without accessory navicular fusion.

|  | Fusion (+) | Fusion (-) | p-value |
| --- | --- | --- | --- |
| Number of feet | 38 | 71 |  |
| Demographic characteristic |  |  |  |
| Sex (male / female) | 21 / 17 | 24 / 47 | 0.03 |
| Age (years) | 12.0 ± 0.7 | 11.9 ± 0.8 | 0.51 |
| Height (cm) | 147.5 ± 8.0 | 151.8 ± 6.5 | 0.003 |
| Weight (kg) | 40.0 ± 8.1 | 45.1 ± 8.4 | 0.003 |
| BMI (kg/m<sup>2</sup>) | 18.3 ± 2.5 | 19.5 ± 3.0 | 0.03 |
| Symptomatic AN (yes / no) | 8 / 30 | 21 / 50 | 0.33 |
| Flatfoot (yes / no) | 12 / 26 | 20 / 51 | 0.71 |
| Sports club participation (yes / no) | 23 / 15 | 50 / 21 | 0.3 |
| Laterality (right / left) | 20 / 18 | 33 / 38 | 0.54 |

Data are presented as mean ± standard deviation or as n (number of feet).
BMI = body mass index; AN; accessory navicular

Table 3 presents the results of the multivariable logistic regression analysis of risk factors for non-fusion of AN. Female sex (odds ratio [OR]: 3.00; 95% confidence interval [CI]: 1.26–7.45; p = 0.01) and higher BMI (OR: 1.20; 95% CI: 1.02–1.43; p = 0.02) were identified as significant risk factors for non-fusion of AN.

**Table 3.**
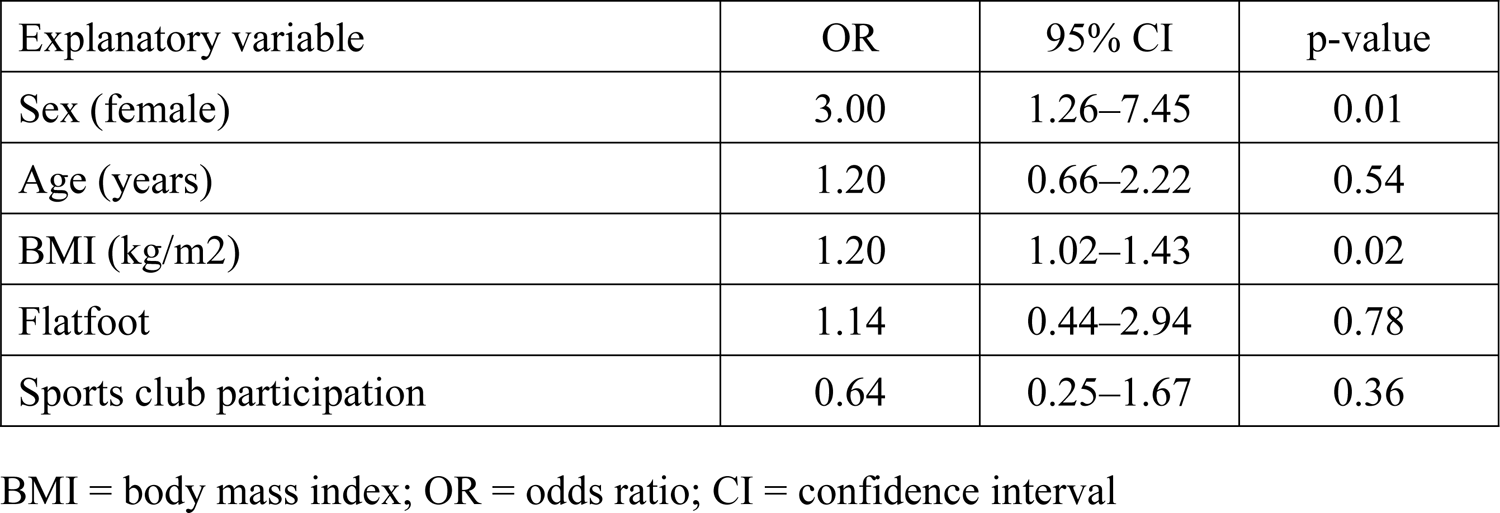
Multivariable logistic regression analysis of risk factors for non-fusion of accessory navicular.

| Explanatory variable | OR | 95% CI | p-value |
| --- | --- | --- | --- |
| Sex (female) | 3.00 | 1.26–7.45 | 0.01 |
| Age (years) | 1.20 | 0.66–2.22 | 0.54 |
| BMI (kg/m<sup>2</sup>) | 1.20 | 1.02–1.43 | 0.02 |
| Flatfoot | 1.14 | 0.44–2.94 | 0.78 |
| Sports club participation | 0.64 | 0.25–1.67 | 0.36 |

BMI = body mass index; OR = odds ratio; CI = confidence interval

## Discussion

The present study investigated the natural history of AN, including symptomatic cases, in Japanese children through a two-year longitudinal analysis of participants aged 11 to 13 years. Findings showed that 34.9% of AN cases underwent fusion with the navicular bone, while pain resolved in 71.4% of those with symptomatic AN. Female sex and higher BMI were identified as independent risk factors for non-fusion of AN. To our knowledge, this is the first study to longitudinally assess the natural course of symptomatic AN. These findings offer fundamental insights that may inform future preventive and therapeutic approaches for managing this condition in pediatric populations.

In the present study, the fusion rate of AN with the navicular bone was 34.9%. To date, only one prior study has reported the fusion rate of AN. Knapik et al. conducted a 13-year longitudinal epidemiological study in the United States, analyzing 261 feet from participants aged 0.25 to 7 years (5). They identified AN in 19 feet, with 8 cases (42%) demonstrating fusion. The reported mean age of fusion was 14.1 ± 2.7 years in males and 12.5 ± 1.0 years in females. The current study, which followed children aged 11 to 13 years over a two-year period, may have underestimated the fusion rate due to the limited follow-up period, as not all participants had reached the fusion age reported in the prior study. Although ethnic background may also influence the results, the findings of Knapik et al. may provide a more precise estimate of fusion rates.

The present study identified female sex as a risk factor for non-fusion of AN. Although no previous research has statistically assessed factors associated with fusion, Knapik et al. reported fusion rates of 33% (4 of 12 feet) in males and 57% (4 of 7 feet) in females in a longitudinal cohort, suggesting a higher rate in females (5). Conversely, our study found fusion rates of 46.7% (21 of 45 feet) in males and 26.6% (17 of 64 feet) in females, indicating a higher fusion rate in males. The prevalence of AN has been reported to vary by ethnicity (24). The study by Knapik et al. was conducted in a cohort of Caucasian children in the United States, whereas the present study involved Japanese children (5). Although it is difficult to make conclusive interpretations regarding the effect of sex on AN fusion, ethnic differences may have influenced the observed variation in fusion rates. Further research is warranted to clarify the impact of sex on AN fusion.

In the present study, higher BMI was identified as a risk factor for the non-fusion of AN. To date, no previous studies have examined the relationship between BMI and AN fusion. Anatomical studies have reported that, in feet without AN, the posterior tibial tendon inserts into the navicular, medial, intermediate, and lateral cuneiforms, the cuboid, and the bases of the second to fourth metatarsals, whereas in feet with AN, the tendon attaches to the AN (25) (26). This anatomical difference suggests that increased BMI may enhance mechanical traction on the AN via the posterior tibial tendon, potentially disrupting the fusion process.

In the present study, 71.4% of children aged 11 to 13 years with symptomatic AN experienced resolution of pain during a two-year follow-up period. To our knowledge, this is the first study to report the natural course of symptomatic AN. Although no previous studies have directly examined the natural history of symptomatic AN, several have reported outcomes of conservative treatment for this condition (27) (28). Wynn et al. reported the outcomes of non-operative treatment in 169 pediatric patients (226 feet) with symptomatic AN, with a mean age of 11.8 years (27). Complete pain relief was achieved in 28% of cases, whereas 30% required surgical intervention. Nguyen et al. evaluated non-operative treatment in children under the age of 19 and reported a success rate of 71.2%, defining treatment failure as the need for surgery (28). They also identified older age as a risk factor for poor outcomes. Considering the findings of the present study alongside previous reports, it may be appropriate to begin with adequate conservative treatment in younger patients with symptomatic AN, while taking into account individual factors such as pain severity and patient preference before considering surgical intervention.

This study has several limitations. First, the cohort included participants aged 11 to 13 years with a two-year follow-up period. This limited range may have resulted in an underestimation of the true AN fusion rate. However, the follow-up period encompassed the age range commonly associated with AN fusion, as reported in a prior study, suggesting that the observed fusion rate likely reflects actual trends. Future studies should incorporate a broader age range and longer follow-up durations. Second, participants were drawn from a single region in Japan, which may limit the applicability of the findings to the national population. Third, all participants were of a single ethnic background, limiting the generalizability of the results to more diverse populations. Fourth, the study did not ascertain whether treatment interventions were provided for symptomatic AN. Thus, it is possible that the pain resolution rate was influenced by such interventions. Notably, no participants underwent surgical treatment. Fifth, AN was not classified using the Veitch classification, which has been shown to categorize many symptomatic AN cases as Type II (4) (29) (30).

## Conclusions

This study delineates the natural history of AN in children, including symptomatic cases, and is the first to report longitudinal data on symptomatic AN. In a two-year longitudinal study of Japanese children enrolled at ages 11 to 13, 34.9% of AN cases fused with the navicular bone, while 71.4% of symptomatic cases showed complete resolution of symptoms. Female sex and higher BMI were associated with an increased risk of non-fusion. These findings may contribute to the foundational understanding necessary for developing targeted prevention and treatment strategies for symptomatic AN in pediatric populations.

## Data Availability

Data cannot be shared publicly because they contain personal information of the research participants. Data are available from the Ethics Committee contact via the Department of Orthopedic Surgery, Wakayama Medical University for researchers who meet the criteria for access to confidential data.

## Acknowledgements

The authors wish to thank Mr. Kenji Oka, Mr. Yasuo Ikeda and Mr. Fumihisa Maeda of the Board of Education in Katsuragi Town, as well as the teachers of the elementary and junior high schools in Katsuragi Town, for their assistance in coordinating the locations and scheduling of participants.

